# Hepatitis C virus infection and tobacco smoking - joint health effects and implications for treatment of both: A systematic review

**DOI:** 10.1101/2021.10.12.21264923

**Authors:** Belaynew W Taye

**Affiliations:** Faculty of Medicine, The University of Queensland; Brisbane, Australia; Mater Research Institute-University of Queensland, Brisbane, Australia; Population Health, QIMR Berghofer Medical Research Institute, Brisbane, Australia; Department of Epidemiology, Bahir Dar University, Bahir Dar, Ethiopia

**Keywords:** smoking cessation, interaction, health effects, anti-HCV therapy, integration

## Abstract

**Background:** Tobacco smoking and hepatitis C virus (HCV) infection cause many diseases independently. The interaction of these conditions on health effects has not been widely studied. There is a paucity of information on addressing tobacco smoking in HCV treatment settings. This review examines the relationship between tobacco smoking and HCV infection and health outcomes and discusses opportunities for treating both conditions.

**Methods:** A systematic review was conducted following the PRISMA 2009 guidelines ***(Registration No.: CRD42019127771)***. We searched PubMed, EMBASE, Web of Science, and CINAHL on the health effects of tobacco smoking and HCV infection using keywords and MeSH terms for hepatitis C, tobacco smoking, hepatocellular carcinoma (HCC), chronic obstructive pulmonary disease (COPD), diabetes mellitus (DM), cardiovascular diseases (CVD), and chronic kidney disease (CKD). We used the Newcastle-Ottawa Scale, a measurement tool to assess systematic reviews (AMSTAR-2), and international narrative systematic assessment (INSA) tools to assess the methodological quality of the included studies.

**Findings:** Tobacco smoking and HCV infection share similar underlying risk factors and hence it is unsurprising that tobacco smoking prevalence is higher in people living with HCV (PLHCV) than in the general population. Tobacco smoking and HCV infection have additive or multiplicative interaction to cause HCC, COPD, DM, CVD, and CKD. Anti-HCV direct-acting antiviral (DAA) treatment is highly efficacious and widely accessible in many countries, but untreated tobacco smoking addiction may undermine the achievement of optimal health outcomes possible from HCV treatment.

**Interpretation:** The scale-up of DAA treatment programs globally is an opportunity to address the high prevalence of tobacco smoking in PLHCV by concurrently offering tobacco smoking cessation treatment. Simultaneous initiation of smoking cessation therapy at HCV treatment centres is likely to be cost-effective at maximizing the health gains afforded by DAA treatment. Studies are needed to evaluate the effect of tobacco smoking cessation on the sustained virologic response in DAA treated patients.

## Introduction

Hepatitis C virus (HCV) is a blood-borne infection belonging to the *Flaviviridae* family of RNA viruses with eight genotypes and over 60 subtypes.^1-3^ Genotype 1 (G1) is the commonest (causing 46% of HCV infections) followed by G3 (22%), G2 (13%), and G4 (13%).^3,4^ Genotypes 5 and 6 are responsible for the remaining <5% of HCV infections.^4^ Global HCV prevalence is estimated at 2.5% (177·5 million cases), with higher rates in European and Eastern Mediterranean regions.^4-6^

The risk factors for HCV infection vary widely from region to region depending on socioeconomic status, behavioural, and healthcare quality dimensions. In the Middle East and Africa, healthcare-associated exposures including blood transfusion, hemodialysis, unsafe medical injections, and surgical and dental procedures are the commonest sources of infection (59·5%) followed by injecting drug use (IDU) (20·9%), community-based exposures (15·8%), and sexual contact (3·7%).^7,8^ IDU is the commonest risk factor for HCV infection in high-income countries (HICs) such as Australia (80%) and the US (32%).^9^ Other risk factors for HCV infection include unsterile tattooing and body piercing, occupational exposure to infected blood, needle-stick injuries, and mother-to-child transmission.^10,11^

Tobacco smoking prevalence is estimated to be 21% globally and tobacco kills more than 7 million people each year.^12^ In low-and-middle-income countries (LMICs), such as those in Latin America (21%), Central, Southern, and Western Asia (23%), and Eastern and South-eastern Asia (23·5%), more than one-in-five adults smoke tobacco.^13^ In most HICs, tobacco smoking prevalence has dropped to well below 20%. For example, current tobacco smoking prevalence among adults was 15·6% in 2016 in Australia^14^, 15·5% in 2016 in the US^15^, and 15·1% in 2017 in the UK.^16^ Despite substantial declines in smoking in the general population, tobacco smoking prevalence remains very high among many sub-populations including people with mental illness^17^, substance use disorders^18^, and HIV.^19^

Although there is a paucity of evidence regarding the prevalence of tobacco smoking in PLHCV, some studies, mostly based on clinical samples, show tobacco smoking is much higher in PLHCV than among the general population. Studies showed prevalence from more than half (52·9%) ^20^ to three-fourths (76·2%)^21^ of PLHCV smoked tobacco.

A number of factors have been associated with tobacco smoking among PLHCV including older age, low education, low income, current use of cannabis, cocaine, heroin, harmful use of alcohol, low self-efficacy, poor social support, perceived benefit of smoking cessation, loneliness, depression and anxiety, pain, and presence of life stressors.^22,23^

Tobacco smoking contributes to premature death and morbidity, mainly from coronary heart disease and stroke, cancers of the lung, upper airways, liver, and stomach, miscarriage and underdevelopment of the foetus.^24^ Tobacco smoking is also a risk factor for chronic kidney disease (CKD), cancer of the colon and pancreas, diabetes mellitus (DM), osteoporosis, cataract and macular degenerations, and dementia.^25^ HCV infection is also a risk factor for many diseases, including hepatocellular carcinoma (HCC), chronic obstructive pulmonary disease (COPD), cardiovascular diseases (CVD), DM, and CKD^26-29^. Despite this, the combined health effects of tobacco smoking and HCV infection have not been well studied at a larger scale as are the treatment opportunities for tobacco addiction in the presence of HCV infection. Co-occurrence of these two risk factors has implications for the treatment of HCV if health outcomes are to be optimized.

DAAs for treatment of HCV demonstrate a high rate of sustained virologic response (SVR) and favourable safety profile compared to previous treatments, such as interferon.^30^ Hence, DAAs are expected to significantly reduce the prevalence of HCV in the coming decades potentially leading to its elimination.^31-33^ While DAA treatment has high efficacy and results in a cure for >95% of those treated, PLHCV often have multiple health issues, including substance use disorders.^22^

This review aimed to describe the interaction between tobacco smoking and chronic HCV infection and the development of chronic health conditions. We reviewed the risk factors for tobacco smoking among PLHCV, potential joint health implications and discussed the opportunities for the treatment of both HCV infection and tobacco smoking addiction.

## Methods

### Search strategy and selection criteria

This systematic review ***(Registration No*.: *CRD42019127771)*** followed the PRISMA 2009 guidelines. We searched PubMed, EMBASE, Web of Science, and CINAHL (until July 31, 2018) for HCV infection, tobacco smoking, and key health effects including HCC, COPD, CVD, DM, and CKD by combining the keywords and MeSH terms for ‘cigarette smoking’, ‘hepatitis C virus’, ‘hepatocellular carcinoma’, ‘chronic obstructive pulmonary disease’, ‘cardiovascular diseases’, ‘diabetes mellitus’, and ‘kidney diseases’.

We included studies that assessed both tobacco smoking and HCV infection and their joint health effects (HCC, COPD, CVD, DM or CKD) in adults. Studies published in a non-English language, conducted in children, and where data on joint health effects of HCV infection and tobacco smoking could not be extracted were excluded.

Initially, all identified records were exported from each database into an EndNote library and duplicate records were removed. The lead author screened the remaining records for relevance to the review by reading the title and abstracts. Then, further screening for eligibility using the above inclusion criteria was made by reading the full text of the articles.

### Risk of bias and quality assessment

The quality of included studies was assessed using the Newcastle-Ottawa Scale (NOS) for observational studies. The rating scale for case-control and cohort designs was used directly^34^ and the modified NOS for cross-sectional studies, as suggested and used by Modesti and colleagues, was used to assess the quality of cross-sectional studies.^35^ We used a measurement tool to assess systematic reviews (AMSTAR-2) tool for assessing the methodological quality of the systematic reviews included.^36^ In this scale, scores of 0–4 indicate low quality, 5–8 indicate moderate quality, and 9–11 indicate high quality.^36^ We assessed the methodological quality of narrative reviews using international narrative systematic assessment (INSA) tool and a score of at least five is considered good for a narrative review assessed by this scale.^37^

### Data extraction and synthesis

We extracted the following information from the final included studies (see tables 1-5): study period, country of study, population type, study setting, sample size, study design, and main outcome measured and discussed the characteristics of included studies under each health effect. The major conclusions that relate to our study objective were synthesized and presented systematically by the results of the interaction of tobacco smoking and HCV infection. We presented HCC, COPD, CVD, DM, CKD and opportunities and benefits of simultaneously treating tobacco smoking and HCV infection.

**Table 1:**
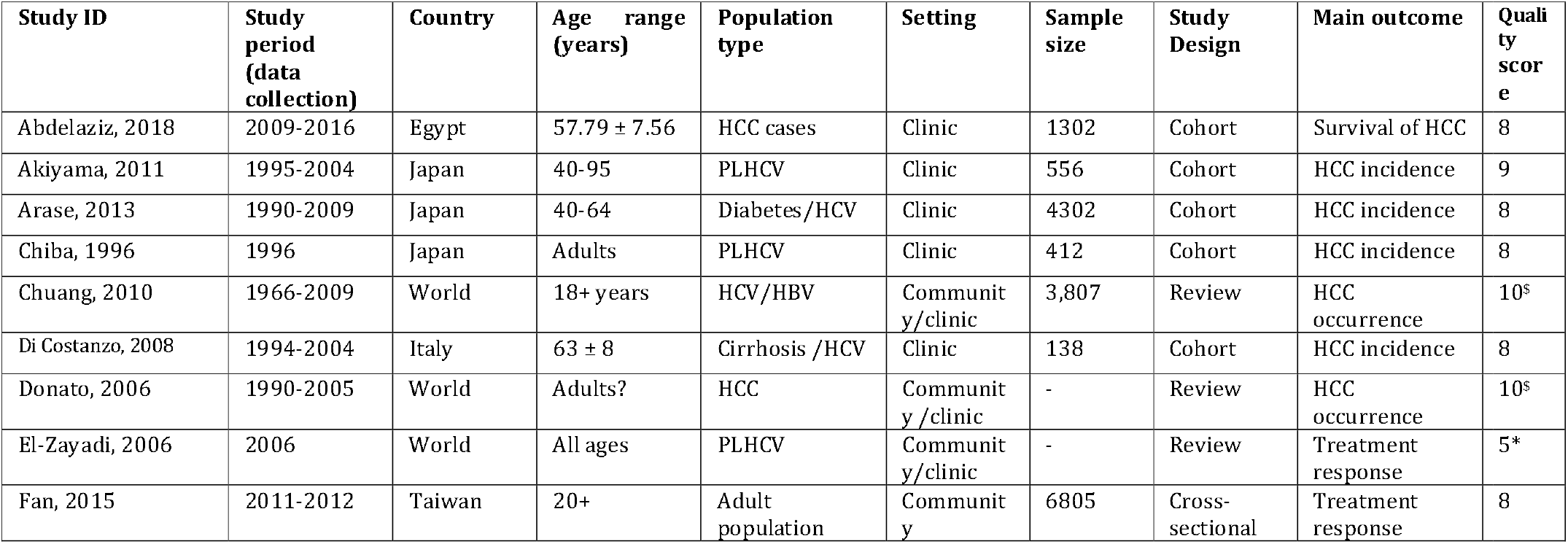
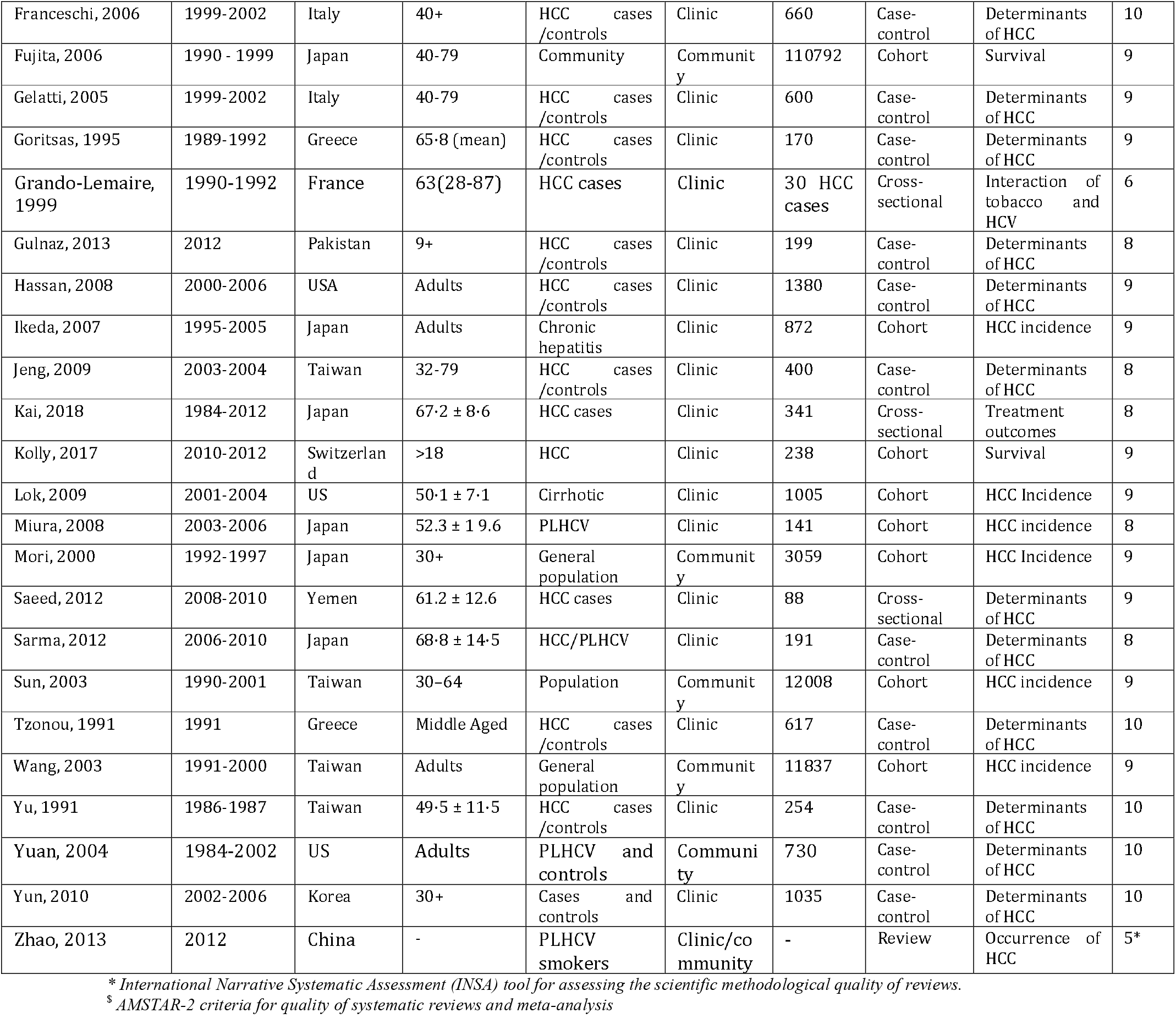
Characteristics of studies assessing the interaction between tobacco smoking and HCV infection on HCC.

**Table 2:** Characteristics of studies on the interaction between tobacco smoking and HCV infection on CVD risk.

| Study ID | Study period (data collection) | Country | Age (years) | Population type | Setting | Sample size | Study Design | Main outcome | Quality score |
| --- | --- | --- | --- | --- | --- | --- | --- | --- | --- |
| Bedimo, 2010 | 1996-2004 | USA | 45.8 ±11.3 | PLHIV /PLHCV | Clinic | 19424 | Cohort | Cerebrovascular events | 9 |
| Cacoub, 2018 | 2006-2015 | France | >18 | PLHCV | Clinic | 878 | Cohort | Major adverse cardiovascular events | 8 |
| Chew, 2015 | 2001-2009 | USA | 20-79 | PLHCV/controls | Community | 237069 | Cohort | Coronary heart disease | 9 |
| Coppo, 2015 | 2005 - 2012 | Italy | >18 | HCV+/- | Clinic | 1266 | Cohort | Microangiopathy | 9 |
| Petta, 2012 | 2012 | Italy | 23-75 | PLHCV /controls | Clinic | 348 | Cross-sectional | Carotid plaques | 9 |
| Roed, 2014 | 2010 - 2011 | Denmark | 51 | PLHCV/controls | Community/clinic | 120 | Cross-sectional | Coronary artery disease | 7 |
| Tsao, 2008 | 2002-2006 | Taiwan | 34.7 ± 13.2 | Transplant | Clinic | 112 | Cohort | Survival | 8 |
| Younossi, 2013 | 1999-2010 | USA |  | General population | Clinic | 19741 | Cross-sectional | Metabolic syndrome | 8 |

**Table 3:** Characteristics of studies on the relationship between tobacco smoking, and HCV infection, and the risk of COPD.

| Study ID | Study period (data collection) | Country | Age (years) | Population type | Setting | Sample size | Study Design | Main outcome | Quality score |
| --- | --- | --- | --- | --- | --- | --- | --- | --- | --- |
| Fischer, 2014 | 2014 | Brazil | >18 | HCV/HIV | Clinic | 1068 | Cross-sectional | COPD | 8 |
| Kanazawa, 2003 | 1997-2002 | Japan | 59.5±6.2 | COPD | Clinic | 59 | Cohort | COPD/FEV1 | 8 |
| Nakashima-2011 | 2004 | Japan | 57.8 | Asthmatic | Clinic | 1327 | Cross-sectional | COPD/Asthma | 8 |

**Table 4:**
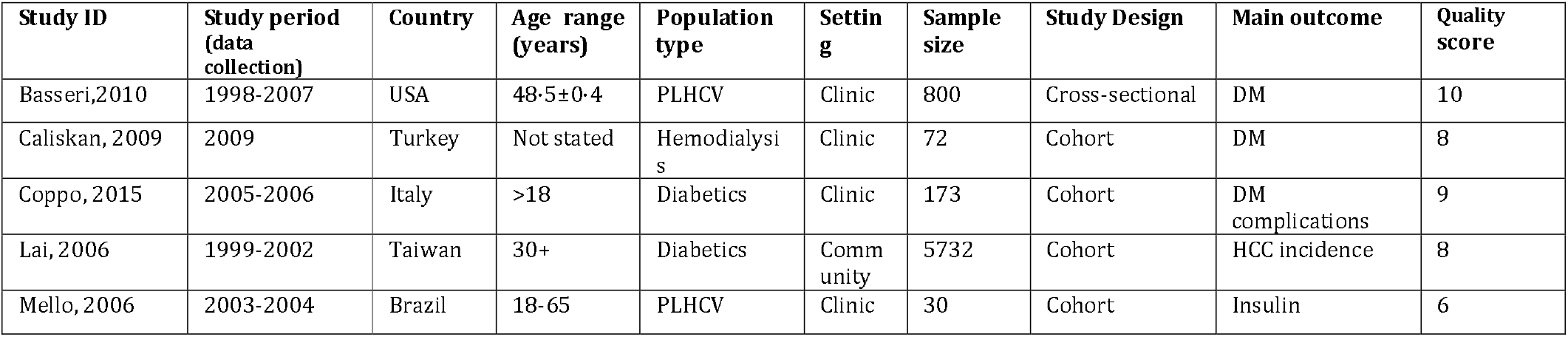
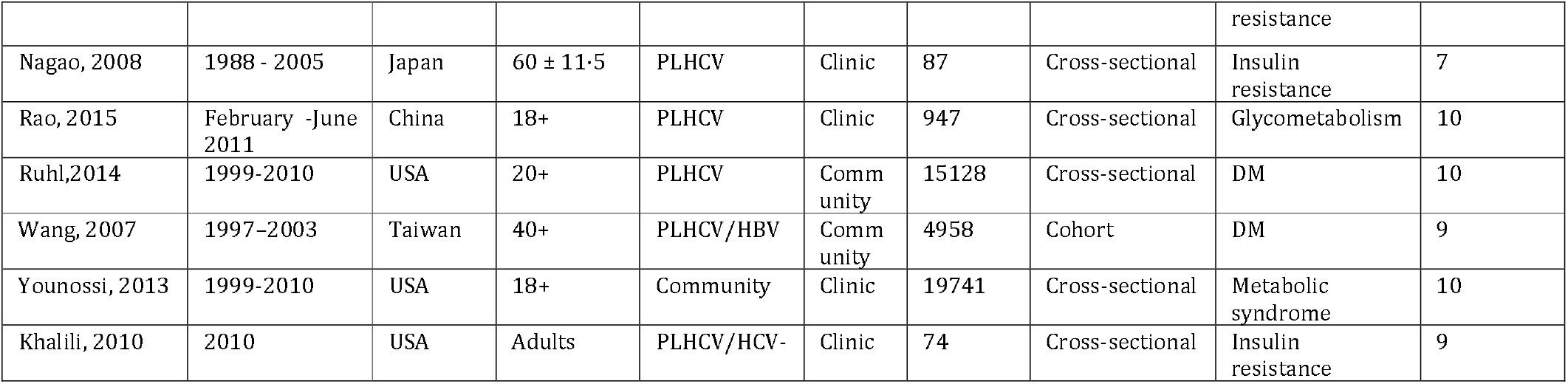
Characteristics of studies on the interaction between tobacco smoking and HCV infection and diabetes mellitus

| Study ID | Study period (data collection) | Country | Age range (years) | Population type | Setting | Sample size | Study Design | Main outcome | Quality score |
| --- | --- | --- | --- | --- | --- | --- | --- | --- | --- |
| Basseri, 2010 | 1998-2007 | USA | 48.5±0.4 | PLHCV | Clinic | 800 | Cross-sectional | DM | 10 |
| Caliskan, 2009 | 2009 | Turkey | Not stated | Hemodialysis | Clinic | 72 | Cohort | DM | 8 |
| Coppo, 2015 | 2005-2006 | Italy | >18 | Diabetics | Clinic | 173 | Cohort | DM complications | 9 |
| Lai, 2006 | 1999-2002 | Taiwan | 30+ | Diabetics | Community | 5732 | Cohort | HCC incidence | 8 |
| Mello, 2006 | 2003-2004 | Brazil | 18-65 | PLHCV | Clinic | 30 | Cohort | Insulin | 6 |
|  |  |  |  |  |  |  |  | resistance |  |
| Nagao, 2008 | 1988 - 2005 | Japan | 60 ± 11.5 | PLHCV | Clinic | 87 | Cross-sectional | Insulin resistance | 7 |
| Rao, 2015 | February -June 2011 | China | 18+ | PLHCV | Clinic | 947 | Cross-sectional | Glycometabolism | 10 |
| Ruhl, 2014 | 1999-2010 | USA | 20+ | PLHCV | Community | 15128 | Cross-sectional | DM | 10 |
| Wang, 2007 | 1997-2003 | Taiwan | 40+ | PLHCV/HBV | Community | 4958 | Cohort | DM | 9 |
| Younossi, 2013 | 1999-2010 | USA | 18+ | Community | Clinic | 19741 | Cross-sectional | Metabolic syndrome | 10 |
| Khalili, 2010 | 2010 | USA | Adults | PLHCV/HCV- | Clinic | 74 | Cross-sectional | Insulin resistance | 9 |

**Table 5:** Characteristics of included studies assessing the interaction between tobacco smoking and HCV infection on chronic kidney diseases.

| Study ID | Study period (data collection) | Country | Age range (years) | Population type | Setting | Sample size | Study Design | Main outcome | Quality score |
| --- | --- | --- | --- | --- | --- | --- | --- | --- | --- |
| Basseri, 2010 | 1998-2007 | USA | 48.5±0.4 | PLHCV | Clinic | 800 | Cross-sectional | Co-morbidities | 10 |
| Butt, 2006 | 1997-1998 | USA | 18-90 | PLHCV/HIV | Clinic | 5737 | Cross-sectional | Co-morbidities | 9 |
| Su, 2015 | 2008-2010 | Taiwan | >18 | PLHCV | Clinic | 10463 | Case-control | Co-morbidities | 9 |
| Underner, 2008 | 2008 | France | Adults | PLHCV | Clinic | - | Review | CKD | 5* |
| Yucel, 2004 | 2004 | Turkey | 25-65 | ESRD | Clinic | 76 | Cross-sectional | Bone mineral density | 7 |

## Results

### Search results and eligibility assessment

Figure 1 presents the PRISMA flowchart for the screening and eligibility assessment of identified records. A total of 464 records were identified. Seventy-four duplicate records were excluded, and we screened 390 records for eligibility. One hundred seventy-five records were excluded by reading the title or abstract.

**Figure 1:**
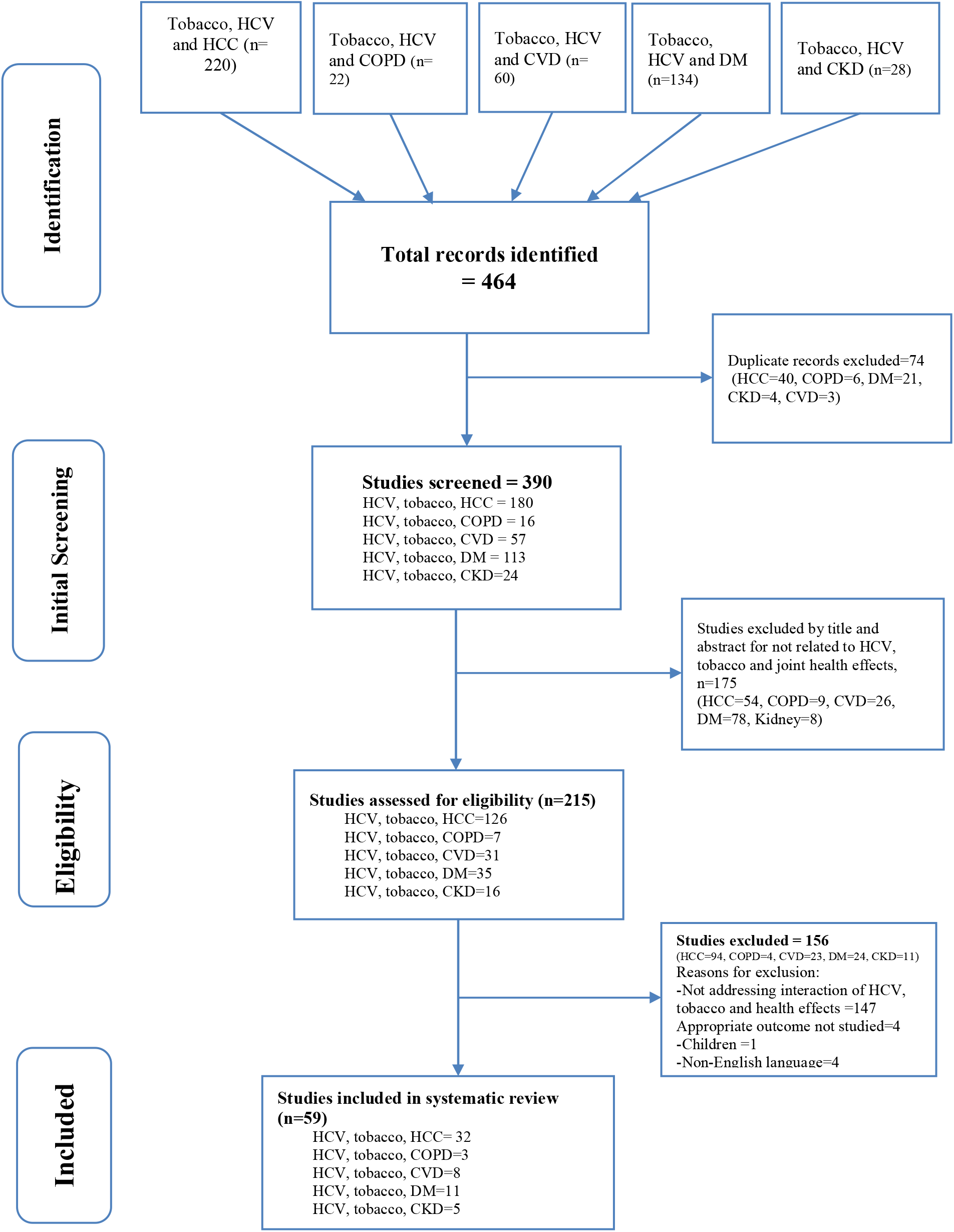
Results of literature search and screening for studies assessing tobacco smoking, HCV infection and the health effects.

We assessed the full text of the remaining 215 studies for eligibility and excluded 156 studies that did not fulfil inclusion criteria. Among these, 147 studies did not address HCV, tobacco smoking and health outcomes and we could not extract data on the interaction effects, one study was in children, an appropriate outcome was not assessed in four studies, and four were non-English language studies (Figure 1). Finally, we included 59 eligible studies in the systematic review.

### Risk of bias and quality assessment in individual studies

We present the quality assessment scores of included studies in tables 1-5 under specific health outcomes. Of seventeen cross-sectional studies included (four studies on HCC, two studies each on COPD and CVD, six studies on DM, and three studies on CKD), nine had a quality score of 9 and above. The NOS scores of other studies were 8 (five studies), 7 (three studies), and 6 (one study). Among 24 cohort studies, 14 had a NOS score of 9, nine studies had a score of 8 and in one study the score was 6. Five out of eleven case-control studies had a NOS score of 10 and the remaining studies had scores of 9 (four studies) and 8 (three studies). Two systematic reviews had an AMSTAR-2 score of 10 and the methodological quality score for each of the three narrative reviews using the INSA scale was 5.

##### Research in context

###### Evidence before this study

Independent health implications of tobacco smoking and hepatitis C virus infection have been stated. The presence of highly efficacious DAA treatment for HCV infection with growing accessibility globally is well documented.

###### Added value of this study

Interaction of tobacco smoking and HCV infection to cause many disease conditions including HCC, COPD, CVD, DM, and CKD. Tobacco smoking impacts adherence to DAA therapy, rate of SVR in interferon-based treated PLHCV, and occurrence of HCC, CVD, DM, and CKD even with good anti-HCV treatment success. The global expansion of DAA treatment leading to a wider access is an opportunity to initiate smoking cessation treatments simultaneously with HCV treatment.

###### Implications of all the available evidence

Simultaneous initiation of tobacco cessation treatment helps to reduce tobacco smoking burden, tobacco related diseases and maximizes the health gains expected from anti HCV therapy.

### Tobacco smoking and chronic HCV infection-joint health implications

Tobacco smoking and chronic HCV infection independently increase the risk of many serious health conditions. However, smoking and HCV infection may also act synergistically for some diseases.

### Hepatocellular carcinoma

Both tobacco smoking (OR = 4·55, 95% confidence interval (CI): 1·90 - 10·91) and chronic HCV infection (OR = 13·36, 95% CI: 4·11 - 43·45) are independently predictive of HCC.^38^ A nested case-control study in Europe that showed the relative importance of tobacco smoking, HCV infection and their unknown interaction effect found 73% of HCC cases were tobacco smokers and nearly one-in-five cases were diagnosed with HCV infection (Figure 2).

**Figure 2:**
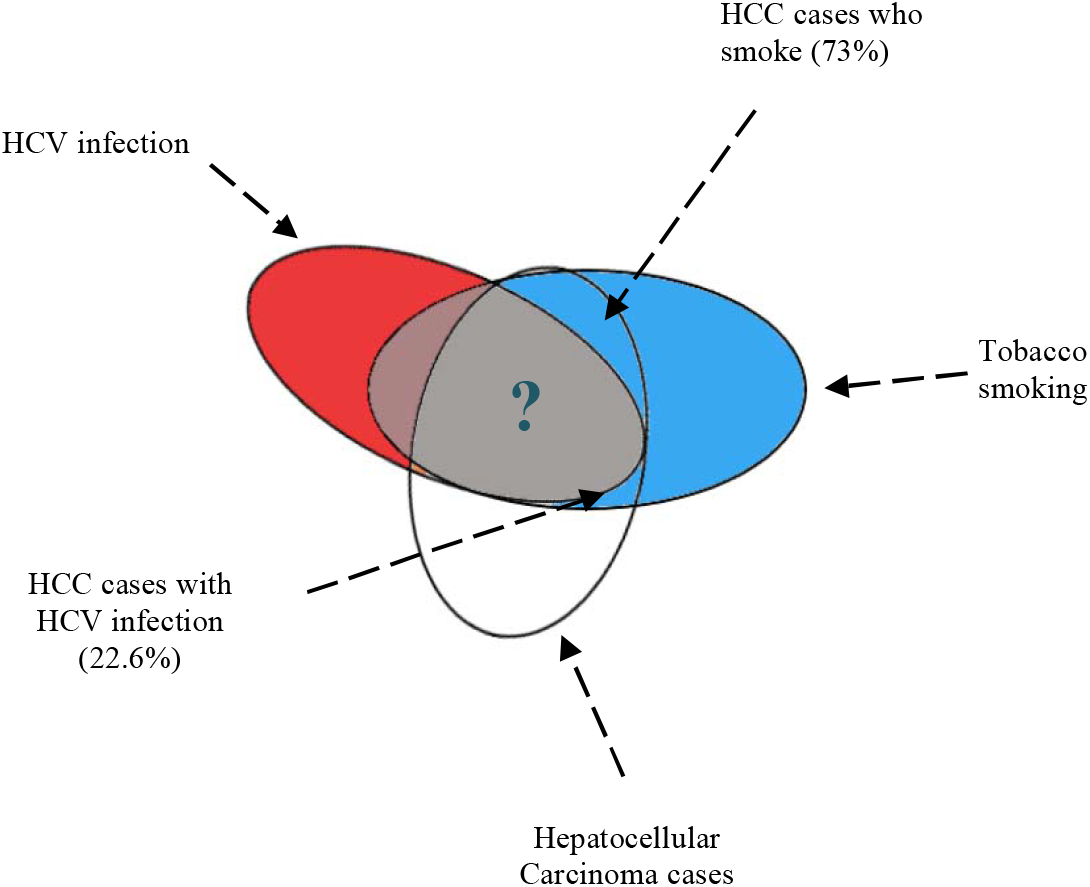
Relative contribution of smoking and hepatitis C virus infection to hepatocellular carcinoma. The grey area shows the unknown proportion of HCC cases who smoke tobacco and diagnosed HCV. **Adapted from (Trichopoulos, et’al, 2011)**

Although the mechanism is not yet well understood, it is hypothesized that tobacco-related carcinogens metabolized by the liver accelerate carcinogenesis caused by chronic HCV infection. Additionally, smoking can contribute to oxidative stress, a weakened immune system, and insulin resistance which may increase the risk of developing HCC due to HCV infection.^39^

Thirty-two studies (13 cohort, 11 case-control, four cross-sectional, and four reviews) assessed the joint effects of tobacco smoking and HCV infection on HCC. Twenty-two of the 32 included studies were clinical whereas six studies were community-based (Table 1).

The risk of developing HCC is higher in PLHCV who smoke compared with non-smokers. A longitudinal clinical study of 4302 adult PLHCV aged 40 years and older in Japan found the hazard of developing HCC in PLHCV who smoked ≥ 20 cig/day was 1·43 times higher compared with PLHCV who smoked < 20cig/day.^40^ A cohort study of HCV related HCC cases in Taiwan found an additive interaction between HCV infection and tobacco smoking on HCC with (RR = 3·9, 95%CI: 2·0 - 7·7) in PLHCV who smoke compared with HCV negative smokers (RR= 1·1, 95% CI: 0·7 - 1·7) and PLHCV who don’t smoke (RR = 2·1, 95%CI: 0·8 - 5·3).^41^

A meta-analysis of six observational studies that calculated the joint effects of tobacco smoking and HCV infection found a multiplicative interaction between the two for HCC risk. The relative risk of HCC was 15·4 times higher for HCV positive smokers compared with HCV negative smokers (RR = 23·1 versus RR=1·5). The effect was nearly 3 times higher for PLHCV smokers compared with PLHCV non-smokers (RR = 23·1 vs RR = 7·94).^39^ In addition, a review of community-based and clinical studies on HCC aetiology in Southern Europe found tobacco smoking as an important co-factor that aggravates the progression of a hepatic lesion and hence the development of HCC in PLHCV.^42^

Tobacco smoking has a negative impact on treatment outcomes of HCV infection. Tobacco smoking in PLHCV has a negative impact on SVR gained from interferon-based therapy and prevention of HCC occurrence.^43,44^ However, the role of tobacco smoking on SVR in DAA treated patients is not yet investigated.

Tobacco smoking increases the odds of dying from HCC in PLHCV. In a case-control study of PLHCV and HCV negative persons in Japan, current smokers had 9·6 times higher odds of dying from HCC (OR = 9·6, 95% CI: 1·50 - 61·36) compared with non-smokers while the odds of death from HCC among HCV negative smokers was 1·71 times higher and not statistically significant (OR = 1·71, 95%CI: 0·58 - 5·08). A dose-response relationship exists between death from HCC mortality and the number of cigarettes smoked per day. PLHCV who smoked ≥20 cig/day had 9·1 times higher odds of dying from HCC (OR = 9·1, 95% CI: 1·10 - 75·05) compared with never-smokers but it was not significant in HCV negative persons who smoked ≥20 cigarettes (AOR = 1·27, 95%CI: 0·38 - 4·20). This study also found the odds of dying from HCC in PLHCV who smoked cigarettes for 35 or more years was nearly 7 times greater than that in never-smokers (AOR = 6·99, 95%CI: 1·03 - 47·51) while it was 1·27 times higher in PLHCV negative persons who smoked for 35 or more years compared with PLHCV negative never-smokers (OR = 1·27, 95%CI: 0·38 - 4·20).^45^

Similarly, in a cohort study of adult HCC patients with HCV/HBV coinfection, smokers had nearly 3 times higher hazards of dying from HCC compared to non-smokers with HCV/HBV infection (HR = 2·99, 95%CI: 1·7 - 5·23).^46^ Both tobacco smoking and HCV infection affect survival after surgery for HCC. In a retrospective cohort study of smoking and surgical outcomes, current smokers with HCV related HCC had 1·69 times higher hazards of dying after hepatic resection compared with non-smokers (HR = 1·69, 95%CI: 1·18 - 2·42).^47^

### Cardiovascular diseases

Eight studies assessed the joint effect of tobacco smoking and HCV infection on CVDs. Five studies employed a cohort design^44,48-50^ while three studies were cross-sectional.^21,26,51^ Six studies were conducted in a clinical setting and two of the eight included studies were community-based. Four studies^44,48,50,51^ had a high NOS quality assessment score of 9, three studies^21,49,52^ had a score of 8, and one study^26^ had a score of 7 (Table 2).

There is a well-documented relationship between HCV infection and tobacco smoking for risk of CVD development.^44,50^ Tobacco smoking in PLHCV contributes to an increased Framingham Risk Score (p<0·001)^50^ and the risk of acquiring major adverse cardiovascular events (HR = 1·86, 95%CI: 1·03 - 3·41).^49,53^ HCV infection and tobacco smoking interact positively to result in increased risk of CVD-related mortality (AOR = 1·65, 95%CI: 1·07 – 2·56), cerebrocardiovascular events (OR = 1·30, 95% CI: 1·10 – 1·55),^52,54^ and the rate of tobacco smoking in the HCV positive cohort, with high rate of cardiovascular diseases, was significantly higher (76·23% vs 29·85%,).^21^

### Chronic obstructive pulmonary disease

It has been theorized that the increased T lymphocytes seen in PLHCV could influence lung function decline in COPD patients.^55^ Furthermore, COPD lung inflammation may be amplified by latent viral infections such as chronic HCV.^56^

Only three studies provided information on the interaction between tobacco smoking, HCV infection, and COPD including asthma (Table 3).

A clinical study of PLHCV and HCV negative adult patients with asthma in an outpatient unit identified that PLHCV with asthma were more likely to be smokers compared with the HCV negative comparison group.^57^ A longitudinal study of 59 COPD patients (15 HCV-negative ex-smokers; 14 HCV-negative current smokers; 14 HCV-positive ex-smokers; 16 HCV-positive current smokers) found the decline in lung function (FEV1) among HCV positive current smokers was higher than ex-smokers or HCV negative current smokers after five years of follow-up. This suggests chronic HCV infection could accelerate a decline in lung function in COPD patients who smoke.^55^ Different from other reports, a clinic-based cross-sectional study of with HCV and/or HIV infection in Brazil found no independent association between HCV and COPD (OR = 0·82, 95% CI: 0·51 – 1·31) and current smoking was not associated with the development of COPD in PLHCV (OR = 1·00, 95% CI: 0·91 – 1·11).^58^

### Diabetes mellitus

Chronic HCV infection is associated with a higher risk of DM as a result of direct viral role or secondary to HCV-induced liver damage, both of which lead to abnormal glucose metabolism.^59^ In PLHCV who smoke, HCV-induced hepatitis, high triglyceride levels, and physical inactivity lead to insulin resistance eventually aggravating the chance of occurrence of DM..^60,61^

Table 4 shows the characteristics of studies that assessed the role of tobacco smoking and HCV infection to cause DM. Three of the 11 studies included were community-based cohort or cross-sectional (Table 4).

The odds of developing DM were 1·4 - 5 times higher in PLHCV who smoke compared with PLHCV who do not smoke. According to a clinic-based cross-sectional study of 800 PLHCV in the US, HCV infection confers 1·44 times higher odds of DM (OR = 1·44, 95%CI: 1·16 - 1·78) compared with general population controls. On the other hand, the odds of tobacco smoking were more than 5 times higher in PLHCV (OR = 5·52, 95%CI: 4·77 – 6·4) compared to HCV negative controls.^62^ Additionally, tobacco smoking in PLHCV is significantly associated with abnormal glycometabolism, insulin resistance, and development of DM.^63-66^ Likewise, current tobacco smoking nearly tripled the hazards of DM-related complications (HR = 2·94, 95% CI: 1·06 - 8·17) compared with never-smokers independent of HCV status, but HCV infection was not independently associated with DM-related complications (HR = 0·74, 95%CI: 0·33 - 1·71).^44^

### Renal diseases

We found five studies that assessed the interaction of tobacco smoking and HCV infection on occurrence and complications of renal diseases (Table 5).

A large cross-sectional study of PLHCV on dialysis found the odds of developing CKD in smokers was 1·59 (95% CI: 1·36 – 187) times higher than that in non-smokers.^67^ In the same way, tobacco smoking in PLHCV increases the risk of renal cell carcinoma and chronic nephropathies.^68^ Tobacco smoking also aggravates the development of bone mineral density defects in PLHCV on haemodialysis.^69^

## Discussion

Tobacco smoking and chronic HCV infection each account for a significant disease burden and mortality worldwide. These two conditions also interact to result in several diseases, including HCC, COPD, CVD, DM, and CKD. Tobacco smoking and HCV infection share a few risk factors that partly explain their co-occurrence and offer the potential to address both health issues simultaneously.

The higher rate of smoking in PLHCV compared with the general population is unsurprising due to their shared underlying risk factors.^20,22^ Even with successful SVR after treatment, tobacco smoking, and HCV infection both contribute to the development of many chronic diseases.^49,70^ This emphasizes that optimal health outcomes expected from DAAs may not be achieved without simultaneously addressing tobacco smoking as many PLHCV will still be at risk of developing many of these health conditions due to their smoking even if they are cured of HCV. Combining smoking cessation assistance with HCV treatment may be a way to optimize health outcomes for this population.

Increased access to highly effective DAAs for the treatment of HCV is revolutionizing the prospect of ending the HCV epidemic. The new DAA therapies have many benefits including improved efficacy to over 90% for all genotypes^9^ compared with only 45 - 75% efficacy for genotypes 1, 2, and 3 with their predecessors.^71^ Several HICs such as Australia, Canada, and the US publicly subsidized HCV treatment to increase coverage of anti-HCV treatment.^9,72,73^ As a result, treatment uptake for HCV significantly increased.^74^ Globally, the number of people who initiated DAA-based treatment for HCV rose between 2015 and 2016, from approximately 1 million to 1·5 million and there is a strategy to increase coverage from 7% in 2015 to 80% by 2030.^75^ However, the success rate of HCV therapy can be undermined by substance use during the treatment period by creating poor treatment adherence and thus relapse of HCV.^76^ Tobacco smoking reduces the rate of SVR in PLHCV treated with interferon-based antivirals but the impact of smoking on SVR in DAAs treated PLHCV is an area of further investigation.

Given that smoking prevalence is high among PLHCV, the importance of reducing smoking in this population should receive greater attention. Therefore, the global initiative to scale-up access to DAAs and the current move to roll-out HCV treatment creates an opportunity for treatment of tobacco addiction among a population with high smoking prevalence.^77,78^ Approaches to address both will have multiple benefits to both patients and the health system and increases the likelihood that DAAs reduce morbidity and mortality rates better than delivering DAAs alone.^79^

Tobacco smokers may face many challenges due to lack of access to care, poor adherence to therapy, and HCV reinfection due to risks such as injecting drug use. However, smokers living with HCV can be assisted to quit smoking with both pharmacotherapy and behavioural interventions where close monitoring of adherence and compliance to treatment is promoted.^80^ This could boost achievement of maximum health benefits from the treatments of both tobacco addiction and HCV infection.^80,81^

Currently, the greatest potential for integrating smoking cessation into HCV treatment is in HICs where both HCV and smoking cessation treatment are often publicly subsidized. However, LMICs present a greater challenge, where rates of diagnosis and treatment of HCV and smoking addiction remain low. As a result, access to effective smoking cessation assistance is limited in many LMICs although WHO includes nicotine replacement therapy on its list of essential medicines. Therefore, country-specific strategy to increase access to both HCV and smoking cessation treatments is vital in LMICs. Furthermore, the lessons from increasing access to HIV treatment could be helpful to improve access to HCV and tobacco addiction. Some options include price subsidy for PLHCV, strengthening the health system, advocacy,^82^ and integrating the smoking cessation services into HCV care settings to provide a one-stop-shop service.

## Conclusions

Tobacco smoking and HCV infection are important global public health concerns each posing numerous health challenges. They also interact in an additive or multiplicative way to result in many serious diseases.

The co-occurrence of tobacco smoking in PLHCV has diverse long-term negative health consequences. This shortens the time to occurrence of health complications such as HCC and increases the risk of occurrence of DM, CVD, CKD, and COPD.

Failing to address tobacco addiction in PLHCV will undermine the potential health gains that could be achieved from highly efficacious DAAs. It is recommended that HCV treatment is used as an entry to provide smoking cessation interventions concurrently. The scale-up of DAAs globally now presents a unique opportunity to address tobacco smoking addiction. Integrated treatment of tobacco smoking at HCV treatment settings can be realized by price subsidy, strengthening the health system, and advocacy.

We recommend studies that evaluate the feasibility and cost-effectiveness of integrating smoking cessation care into HCV treatment settings, particularly in LMICs. Population-based studies evaluating the confounding effect of alcohol on the interaction between tobacco smoking and HCV infection are required. We also recommend longitudinal studies to investigate the effect of tobacco smoking on SVR in PLHCV treated with DAAs.

## Data Availability

All data produced in the present work are contained in the manuscript.

## Acknowledgements

We thank the Herston Health Sciences Library for technical support during literature searching.

